# Pathway of Low Anterior Resection Syndrome (LARS) Relief After Surgery (POLARiS) Trial Protocol A prospective, international, open-label, multi-arm, phase 3 randomised superiority trial within a cohort, with economic evaluation, process evaluation and qualitative sub-study, to explore the natural history of Low Anterior Resection Syndrome (LARS) and compare trans-anal irrigation and sacral neuromodulation to optimised conservative management for people with major LARS following a high or low anterior resection for colorectal cancer

**DOI:** 10.1101/2024.08.19.24312209

**Authors:** Julie Croft, Emily Farrow, Alexandra Harriet Coxon-Meggy, Katie Gordon, Neil Corrigan, Hannah Mather, Deborah Stocken, Megan Dale, Huey Yi Chong, Judith White, Laura Knight, Alun Meggy, Christina Lloydwin, Betty Tan, Ashley Douglas, Ralph Powell, Julie Hepburn, David Jayne, Jared Torkington, Andrea Warwick, Kheng-Seong Ng, Kate Wilson, Charles Knowles, Aaron Quyn, Julie Cornish

## Abstract

**Introduction:** As a result of improving survival rates, the adverse consequences of rectal cancer surgery are becoming increasingly recognised. Low Anterior Resection Syndrome (LARS) is one such consequence and describes a constellation of bowel symptoms after rectal cancer surgery which includes urgency, faecal incontinence, stool clustering and incomplete evacuation. LARS has a significant adverse impact on Quality-of-Life (QoL) and symptoms are present in up to 75% of patients in the first year after surgery. Despite this, little is known about the natural history and there is poor evidence to support current treatment options.

**Methods and Analysis:** The objectives of POLARiS are to explore the natural history of LARS and to evaluate the clinical and cost-effectiveness of trans-anal irrigation (TAI) or sacral neural modulation (SNM) compared to optimised conservative management (OCM) for people with major LARS.

POLARiS is a prospective, international, open-label, multi-arm, phase 3 randomised superiority trial within a cohort (TWiCs design), with internal pilot phase, qualitative sub-study, process evaluation, and economic evaluation. Approximately 1500 adult participants from UK hospitals and 500 from Australian hospitals who have undergone a high or low anterior resection for colorectal cancer in the last 10 years will be recruited into the cohort. 600 participants from the UK and 200 participants from Australia, with major LARS symptoms, defined as a LARS score of ≥30, will be recruited to the randomised controlled trial (RCT) element. Participants entering the RCT will be randomised between OCM, TAI or SNM, all with equal allocation ratios.

Cohort and RCT participants will be followed up for a 24-month period, completing a series of questionnaires measuring LARS symptoms and QoL, as well as clinical review for those in the RCT. A process evaluation, qualitative sub-study and economic evaluation will also be conducted.

The primary outcome measure of the POLARiS cohort and RCT is the LARS score up to 24 months post registration/randomisation. Analyses of the RCT will be conducted on an intention-to-treat basis. Comparative effectiveness analyses for each endpoint will consist of two pairwise treatment comparisons: TAI vs OCM and SNM vs OCM. Secondary outcomes include health-related QoL, adverse events, treatment compliance and cost effectiveness (up to 24 months post registration/randomisation)

**Ethics and Dissemination:** Ethical approval has been granted by Wales REC 4 (reference: 23/WA/0171) in the UK and Sydney Local Health District HREC (reference: 2023/ETH00749) in Australia. The results of this trial will be disseminated to participants upon request and published on completion of the trial in a peer-reviewed journal and at international conferences

**Trial Registration Number:** ISRCTN12834598

Registered 04/08/2023

ACTRN12623001166662

Registered 10/11/2023

**Strengths and Limitations:**

- The trial is pragmatically designed to optimise and assess recruitment and retainment.
- This trial includes an economic evaluation of treatment options specific to both the UK and Australia.
- Lay representatives with personal experience of bowel cancer and LARS have contributed throughout the trial design and ongoing Trial Management Group meetings.
- There are recognised potential limitations to the LARS score, including limited sensitivity to detect real time change in response to treatment. Additional outcome measures of Quality of Life and a new LARS Patient Reported Outcome Measure (PROM) are being collected to give a more nuanced picture of treatment response.

## Introduction

Colorectal cancer is the fourth most common cancer in the UK, with an average of 44,100 new cases diagnosed per year. Of these, around 30% of patients will have rectal cancer, making it the most common site of colorectal cancer(1). Many of these patients will undergo major resection(2). With evolving surgical technique and development of adjuvant and neoadjuvant treatments the 5-year survival rate for colorectal cancer has improved by around 30% over 4 decades(3).

As a result of improving survival rates, the adverse consequences of rectal cancer surgery are becoming increasingly recognised. Low Anterior Resection Syndrome (LARS), initially defined in 2012, is one such consequence and describes a constellation of bowel symptoms after rectal cancer surgery which includes urgency, faecal incontinence, stool clustering and incomplete evacuation(4). LARS has a significant adverse impact on Quality-of-Life (QoL)(5). LARS symptoms are present in up to 75% of patients in the first year after surgery, remaining in up to half of these patients over 10 years(6,7). Due to the significant burden and persistence of LARS symptoms, there is an urgent need to improve the understanding around the natural history and treatment for LARS.

Conservative management of LARS includes dietary modifications, medication and physiotherapy. Dietary modifications are hugely variable with many patients implementing their own trial and error strategies(8). Input from dieticians may help to meet this challenge, but often these services are not readily available(9). Pelvic floor physiotherapy and biofeedback techniques have been reported to improve symptoms, although only a few low powered studies have been published(10). These services are not readily available at all hospitals and can have prolonged waiting times(11). Overall, there is little structured guidance on conservative management with a high degree of variability in what centres offer. Conservative treatment for LARS largely targets faecal incontinence symptoms, however, many patients present with predominantly obstructive symptoms such as emptying difficulties and tenesmus(12)(13). Little is known about whether there is a difference in treatment efficacy in patients presenting with a different cluster of symptoms.

For those patients who fail conservative management or suffer from major LARS (LARS score ≥30), treatment options include trans-anal irrigation (TAI) and sacral neuromodulation (SNM). Several small RCTs and studies have assessed the efficacy of TAI on bowel function after low anterior resection, but the sample sizes are small with variable uptake and significant heterogeneity (14)(15)(16). There is variable use of TAI following surgery and the timing, with less than a third of surgeons considering TAI as an option for the treatment of LARS(17). Currently, SNM is only licenced for use in faecal incontinence, but a recent meta-analysis of 10 studies suggests that significant improvements in function might be achieved in LARS, however these conclusions are limited by small sample size and significant heterogeneity between studies(18). Most recently, the SANLARS RCT showed both symptom and quality of life improvement at 6 and 12 month follow up in patients with major LARS(19).

As patients are increasingly surviving rectal cancer surgery it is essential we examine the impact and consequences on patient’s lives and define treatment pathways which are currently lacking robust evidence from RCTs. The National Institute for Clinical Excellence (NICE) have developed a research recommendation to determine the effectiveness and safety of SNM and TAI compared to symptomatic treatment for people with major LARS(20). As very few studies report beyond 12 months, we also lack knowledge on the natural history of LARS. Is there a time-point where LARS symptoms become stable? If we delay treatment post-operatively are symptoms managed as effectively? Is there a difference in long-term outcomes in patients who have had radiotherapy? Understanding these may allow us to make more informed decisions regarding the initiation of treatment and be able to better advise patients on how their symptoms and treatment may change over time. POLARiS is a large-scale, complex randomised superiority trial that will address these research priorities.

## Objectives

- The primary objective of the POLARiS cohort is to explore the natural history of LARS over time.
- The primary objective of the POLARiS RCT is to evaluate the clinical and cost-effectiveness of TAI or SNM compared to OCM for people with major LARS.

## Methods & Analysis

### Study Design

POLARiS is a prospective, international, open-label, multi-arm, phase 3 randomised superiority trial within a cohort (TWiC), with internal pilot phase, qualitative sub-study process and economic evaluation (Figure 1)(21). The Clinical Trials Research Unit (CTRU) at the University of Leeds will perform co-ordination of the trial with Cardiff & Vale University Health Board (C&V UHB) as UK sponsor and University of Sydney as the Australian sponsor.

Patients, aged ≥ 18 years who have had a high or low anterior resection within 10 years for rectal cancer with a functioning anastomosis and meet the eligibility criteria will be invited to the cohort (Table 1). Patients with major LARS and who meet the eligibility criteria will be invited to take part in the RCT. Participants in the cohort and RCT will be asked to complete a set of baseline questionnaires. At 3-monthly intervals following registration/ randomisation participants will complete a LARS score and a series of QoL Questionnaires. The RCT treatments are optimised conservative management (OCM), TAI and SNM. Neither clinicians nor participants will be blinded to the RCT treatment allocation.

**Table 1.** Eligibility criteria for both cohort and RCT participants.

|  | **Cohort Eligibility Criteria** | **RCT Eligibility Criteria** |
| --- | --- | --- |
| **Inclusion Criteria** | • Aged ≥ 18 years<br>• Able to provide written informed consent<br>• Diagnosis of rectal or sigmoid cancer<br>• Low or high anterior resection (colorectal resection with anastomosis to the rectum)<br>• Functioning anastomosis<br>• Primary surgery less than 10 years before recruitment<br>• At least 6 months since reversal of stoma or primary surgery if no stoma created<br>• Able and willing to comply with the terms of the protocol including participant completed questionnaires | • Aged ≥ 18 years<br>• Able to provide written informed consent<br>• Diagnosis of rectal or sigmoid cancer<br>• Low or high anterior resection (colorectal resection with anastomosis to the rectum)<br>• Functioning anastomosis<br>• Primary surgery less than 10 years before recruitment<br>• At least 6 months since reversal of stoma or primary surgery if no stoma created<br>• Able and willing to comply with the terms of the protocol including participant completed questionnaires<br>• Major LARS symptoms within the last 3 months (defined as a LARS score of $\geq 30$)<br>• Clinically appropriate for randomisation as determined by treating clinician |
| **Exclusion Criteria** | • Receiving ongoing chemotherapy, radiotherapy or immunotherapy treatment for cancer<br>• Anterior exenteration | • Receiving ongoing chemotherapy, radiotherapy or immunotherapy treatment for cancer<br>• Metastatic disease<br>• Inflammatory bowel disease<br>• Pregnancy<br>• Use of TAI for LARS within 1 month prior to randomisation<br>• Not eligible for SNM **and** not eligible for TAI<br>• Anterior exenteration<br>• Anastomotic stricture<br>• History of anastomotic leak with evidence of ongoing leak/sinus<br>**Exclusion criteria for SNM:**<br>• Site unable to offer SNM as a treatment<br>• Previous SNM<br>• Margin Positive (R1) resection within 24 months prior to randomisation<br>• Specific contraindications to implantation<br>• Any other contraindications advised by the care team, product manufacturer or distributor<br>**Exclusion criteria for TAI:**<br>• Unable to perform TAI<br>• Previous use of TAI for LARS<br>• Site unable to offer TAI as a treatment<br>• Any other contraindications advised by the care team, product manufacturer or distributor |

### Study Population

1500 UK participants and 500 Australian participants will be entered into the cohort with the aim to randomise 600 from the UK and 200 from Australia into the RCT. Each site must fulfil a set of pre-specified criteria, and the local Principal Investigator (PI) must complete a form to verify the site is willing and able to comply with trial requirements. The trial will open in at least 20 UK and 15 Australian hospitals.

Participating sites must be able to offer at least one of the interventions (TAI or SNM), perform at least 30 anterior resections for colorectal cancer each year and at least 10 SNM procedures each year if delivering SNM to trial participants. Sites which offer TAI and have an existing Service Level Agreement (SLA) with a centre offering SNM, have the option for three-way randomisation.

### Recruitment

Patients will be screened through cancer databases, clinical notes and clinics at participating hospital sites. Those who meet the cohort eligibility criteria (Table 1) will be provided with a cohort participant information sheet (PIS). Once written informed consent is obtained, participants are registered using a central web-based 24-hour registration system (Supplementary 1. Cohort Consent Form).

Participants identified as having a major LARS score (LARS score ≥30), documented within the last three months, at the point of screening and who meet the RCT eligibility criteria (Table 1) can be considered for direct entry into the RCT without registration into the cohort. Participants from the cohort who subsequently become eligible for the RCT (e.g. identification of a major LARS score through completion of the cohort LARS questionnaire) will be invited to participate in the RCT. Eligible patients will be provided with the RCT PIS and will discuss the trial with a suitably qualified member of the health care team. Participants will be given as much time as required (ideally a minimum of 24 hours) to consider the information before written informed consent is taken. Randomisation will take place as soon as possible after written informed consent is obtained (Supplementary 1. RCT Consent Form).

### Randomisation

There are three randomisation options, dependent on the patient’s eligibility (Table 1) and the site’s capacity to deliver TAI or SNM, each with equal allocation ratios, listed below:

1. OCM vs TAI vs SNM
2. OCM vs SNM
3. OCM vs TAI

Randomisation will be performed using the central automated web-based 24-hour randomisation system. Within each randomisation option a computer-generated minimisation programme incorporating a random element (22)(23)will be used to allocate patients with a 1:1(:1) ratio, with the following minimisation factors: recruiting site, time from surgery, biological sex at birth, age, radiotherapy and procedure (high or low anterior resection).

Participants who were initially eligible for and randomised via the 3-way randomisation option, may be eligible for second randomisation if they do not respond to their first allocated treatment option. For participants who undergo a second randomisation, data will continue to be collected in line with primary randomisation timelines.

### Patient and Public Involvement

A total of four lay representatives from the UK & Australia with relevant personal experience of colorectal cancer, LARS and SNM were recruited. Lay representatives contributed to trial development, production of the lay summary and have made active contributions in co-applicant group meetings to guide decisions and provide recommendations.

An additional focus group was conducted in June 2021 with 12 bowel cancer patients from the UK & Australia to investigate key aspects of the trial design covering key areas such as acceptability of randomisation and treatments. This resulted in the development of the three randomisation options, the option of a second randomisation and adjustments to the OCM pathway.

## Interventions

All RCT patients will be provided with a LARS Information Booklet, a detailed support document developed by experts specifically developed for use in the POLARiS trial. The booklet aims to inform and advise participants regarding their condition.

### OCM

The OCM package has been designed by pelvic floor and LARS experts based on current evidence and reviewed by PPI representatives(24). Sites will be trained to deliver the OCM through Site Initiation Visits, a POLARiS specific training video and support document for healthcare professionals. The OCM package includes an initial appointment to establish which symptoms are most bothersome and the impact this has on the participants daily activities, followed by the provision of practical support and advice as well as tailored treatment options such as diet, medication and physiotherapy.

Practical support includes toilet access schemes, toilet positioning, bowel habit training and recommending support groups. Dietary advice includes completion of a food diary and onward referral to a dietician if available at that site. The clinician also completes a review of the current medication and will discuss medication options. All patients are encouraged to complete pelvic floor exercises and onward referral to a pelvic floor physiotherapist is recommended if available at that site.

An additional POLARiS specific OCM support document summarising the above information is provided to all participants randomised to OCM. Onward referral can be made to a dietician and pelvic floor physiotherapist if available at that site. Further advice and medication adjustments will be offered during the 3-month post randomisation visit (Figure 3).

The following interventions used in this study are CE and/or UKCA marked and being used within their licensing specification. Due to the nature of the interventions, this is a non-blinded trial.

### TAI

TAI involves instilling warm water into the rectum and colon via the anus to empty out the stool. There are several commercially available TAI systems and broadly, systems can be divided into low and high-volume. TAI can be delivered via a variety of different systems the choice of which is determined by a clinician. Each site will be responsible for the procurement of irrigation systems. Treatment involves a one-hour education session with a nurse and provision of a starter pack. Participants will have a follow up/troubleshooting review 1-month after the education session (Figure 4).

### SNM

Participants randomised to SNM will undergo a two-stage procedure. Initially a temporary device is fitted with a 2-week test phase. The response is assessed during the test phase and if deemed successful by the patient and clinician a permanent device is fitted (Figure 5). Both procedures are performed as day-cases. Post-operative care will be as per each site’s standard practice. The decision of which SNM device is used, and whether to use a temporary wire electrode or the tined quadripolar electrode lead for the test phase will be at the discretion of the clinical team/patient.

### Follow Up

Cohort follow-up is over a 24-month period. Demographic information and relevant medical history will be collected at baseline for all cohort participants. Follow-up data will be collected from medical notes at 12 and 24-months post registration. In addition, cohort patients will be asked to complete the LARS score on a 3-monthly basis throughout the 24-month follow up period and a set of questionnaires (LARS iCAT, EORTC QLQ-C29 and EORTC QLQ-CR30) at baseline and at 3-, 6-, 12- and 24-months post-registration (Figure 2).

RCT participants will complete the same set of questionnaires at the same time intervals as the cohort participants, with the addition of EQ-5D-5L and Health Resource Use questionnaires. They will also have a series of clinical reviews at baseline and at 3-, 6-, 12- and 24-months post-randomisation in person, via telephone and clinical notes with some variation depending on which treatment the participant has been randomised to (Figure 3, 4 and 5). Participants for both cohort and RCT can choose to complete their questionnaires electronically or on paper. Should a participant be recruited to the RCT from the cohort, the cohort follow-up will cease, and the participant will be follow-up as per the RCT timelines only.

### Sample Size Calculation

The minimal clinical important difference (MCID) is defined as a 5-point difference in LARS score(25). A conservative standard deviation in LARS scores of 15 is assumed(26). The primary outcome is LARS score over 24 months post-randomisation adjusting for baseline. The required sample size for the adjusted analysis is obtained by multiplying the sample size requirement for the unadjusted analysis by a factor of (1-r^2), where r is the correlation between the 24-month and baseline scores[30], assumed to be a weak correlation of 0.3. Attrition is assumed to be no more than 15%.

The number of patients recruited through three randomisation options: i) SNM vs TAI vs OCM ii) SNM vs OCM iii) TAI vs OCM are denoted n1, n2 and n3 respectively hence (2/3)*n1 + n2 patients and (2/3)*n1 + n3 patients will contribute to the SNM vs OCM and TAI vs OCM comparisons respectively. It is assumed that 50% of RCT participants will be entered via randomisation option i), 25% via randomisation option ii), and 25% via randomisation option iii) hence for a total of N participants, 0.25*N + (2/3)*0.5*N will contribute to each of the two primary comparisons.

326 participants are sufficient to yield 85% power to detect the MCID in an (unadjusted) two-sided t-test of 24-month LARS scores at the 5% level of significance (nQuery v3.0). Applying baseline adjustment factor of (1-0.3^2)=0.91 and accounting for 15% attrition results in a sample size target of 350 participants for each comparison. Recruiting a total of 600 participants to RCT allows 350 patients needed for each primary comparison (0.25*600 + (2/3)*0.5*600). Including an additional 200 Australian participants inflates the power to over 90% power to detect the MCID.

## Outcome Measures

### Primary outcome measure (RCT and cohort study)

The primary outcome measure for both the cohort study and the RCT is the LARS score over 24 months. The LARS score is an internationally validated five-question assessment exploring different bowel dysfunction and their frequency (27). The overall score (maximum 42) corresponds to either no LARS (0–20), minor LARS (21–29) or major LARS (≥30). The LARS score will be collected at baseline and 3-monthly intervals until 24-months post registration to cohort or randomisation.

### Secondary outcome measures (RCT and cohort study)

Health-related quality of life will be measured using the EORTC QLQ – C 29 and EORTC QLQ – CR 30, both internationally validated, cancer specific quality of life questionnaires covering multiple domains including emotional and physical function (28) (29). The EORTC QLQ – CR29 specifically focuses on colorectal cancer.

The LARS iCAT (Impact and Consequences Tool) is a new patient reported outcome measure (PROM) developed by an international group. The tool is based on the international consensus definition of LARS, involving a large international patient panel, which identified eight symptoms and eight consequences of LARS(12). LARS iCAT is intended as comprehensive tool to assess treatment response and enable patient phenotyping. The LARS iCAT will be collected as part of the questionnaire set with the aim of validating the tool prior to use in clinical practice.

Information on all adverse events with a causal relationship to the trial or trial interventions will be collected for this trial whether volunteered by the participant, or detected by investigator on questioning, physical examination or other investigations. This is for both cohort and RCT patients from the date of registration/ randomisation throughout the 24-month follow-up period. Adverse events will be graded for severity.

### Secondary outcome measures (RCT only)

Treatment compliance will be measured via appointment attendance, ongoing use of TAI and decision to have the permanent SNM device fitted. An additional quality of life tool, the EQ-5D-5L, will be used to generate a single index value for health status which is valuable in the assessment of health care evaluation and economic analysis(30). A Health Resource Use Questionnaire is also included for the health economics assessment.

### Data Analysis Plan

All analyses will be pre-specified in a Statistical Analysis Plan according to published guidance(31). Analysis of the cohort data will include exploratory model-fitting to evaluate the longitudinal trends in LARS scores in relation to patient characteristics and clinical data, as well as the identification of potential risk factors for major LARS. Analyses of the RCT will be conducted on an intention-to-treat basis. Comparative effectiveness analyses for each endpoint will consist of two pairwise treatment comparisons: TAI vs OCM and SNM vs OCM. Treatment groups will be combined across different randomisation options for gains in efficiency(32). There is no interim analysis planned.

### Primary Analysis

For each of the pairwise comparisons (SNM vs OCM, and TAI vs OCM) the primary analysis plan is to estimate the expected difference in LARS scores at 24 months post-randomisation, using constrained longitudinal multi-level statistical models(33). Transformations of the LARS score and/or non-Normal error assumptions will be considered as required based on the observed distribution of scores. The stratification factors “time between anterior resection and randomisation”, “biological sex at birth”, “age”, “radiotherapy” and “procedure” will be included as fixed effects in the model.

The model will account for nesting of repeated LARS score observations within patient, and the nesting of patients within “Centre” within “Country” by using appropriate variance components to model random effects(34). Randomisation option will also be accounted for in the model (35). Point estimates of treatment effects will be reported with 2-sided 95% confidence intervals.

Missing data mechanisms will be explored. Multiple Imputation using Chained Equations (MICE) will be considered to explore the potential impact of missing data under a MAR assumption, and to explore sensitivity of the results to MNAR mechanisms as required.

### Secondary Analysis

For all continuous secondary endpoints two pairwise comparisons will be made (TAI:OCM, SNM:OCM), with analysis populations and modelling approaches as already described for the primary outcome measure. Transformations of the outcomes and/or non-Normal error assumptions will be considered as required based on the observed distribution of outcomes. Questionnaire domains will be scored according to scoring manuals and reported graphically over time. Adverse events will be reported descriptively.

## Qualitative Sub-Study, Process and Economic Evaluation

The qualitative sub-study, process and economic evaluation will be conducted as separate components in each country to reflect local differences. The Centre for Healthcare Evaluation, Device Assessment and Research (CEDAR) at C&V UHB are responsible for the design, implementation, management and analysis of the UK economic evaluation, process evaluation and qualitative sub-study. The NHMRC Clinical Trials Centre (CTC) at the University of Sydney will undertake an Australian health system cost effectiveness evaluation. The Australian qualitative sub-study will be undertaken by the Surgical Outcome Research Centre (SOuRCe) at Royal Prince Alfred Hospital and the University of Sydney.

### Qualitative Sub-Study

Few qualitative studies have explored patient perspectives on LARS treatments strategies (36). The purpose of the qualitative sub-study is to explore the impact of the trial interventions on participants’ i) quality of life; ii) activities of daily living; iii) LARS symptoms; iv) psychological functioning. Participants who are randomised into the RCT will be given the opportunity to take part in the qualitative sub-study.

In the UK a series of 3 semi-structured interviews will take place over the 24 months follow-up period. In Australia the 3 interviews will take place over 12 months. Demographic data will be used to purposively sample a broad range of participants. Up to 48 UK participants and 24 Australian participants will be interviewed. An interview topic guide will be formulated with input from PPI representatives and healthcare professionals (Supplementary 2 & 3). Interviews will be carried out by a qualitative researcher who is independent of the participants care team.

Transcripts will be imported and coded using NVivo (QSR International). Analysis of the qualitative data will use a mainly iterative-inductive approach and further line by line coding will allow for identification of emerging themes. Results from this will then be integrated with the results from the main trial to fully and accurately reflect patient experiences of LARS and the impact of the trial treatments. Data from Australian interviews will form a stand-alone publication.

### Process Evaluation

The purpose of the process evaluation is to provide context to the efficacy findings of the study. It will use mixed methods to investigate the domains of acceptability, implementation, including fidelity, dose and reach, mechanisms of impact and context, as defined by MRC guidance for process evaluations(37). A mixed-methods approach will be taken using data from patient surveys (optional for patients, administered at 3-month and 24-month follow ups), staff interviews (n=24 from 12 sites during the first and last six months of recruitment), study records and clinical case report forms (CRFs). Staff interviews will be audio-recorded and used to create anonymised transcripts. These transcripts and qualitative data from survey responses will be analysed thematically using NVivo (QSR International) (38). Quantitative data will be analysed descriptively using a statistical package (SPSS 29). Qualitative and quantitative findings will be integrated at the conclusion of the study.

A process evaluation pilot report, using the pilot patient survey and staff interview data will be presented to the TMF following the internal pilot to facilitate the optimisation of study procedures for participants and staff.

### Economic Evaluation

The economic evaluation will include a within-trial economic analysis of patient-level trial data and a long-term decision analytic model to extrapolate long-term costs and outcomes of each treatment group. In both the UK and Australia, RCT participants will complete a Health Resource Use Questionnaire at baseline, 3-, 6-, 12- and 24-months post-randomisation. This will capture data such as primary, secondary and private health care resource use, personal expenses on consumables, medicines and travel costs. In Australia, RCT participants’ use of medical services and prescription medicine in the primary care setting will be linked to Medicare and Pharmaceuticals Benefits data with Services Australia. CRFs will be used to capture patient-level resource use associated with the treatment during the trial period.

Effectiveness will be measured in terms of quality-adjusted life years. The primary analysis will evaluate cost-effectiveness from a health care payer perspective and a secondary analysis will be undertaken to include wider societal costs. Deterministic and probabilistic sensitivity analyses will be performed to characterise the parametric uncertainty associated with the cost and outcome differences between groups. The reporting of this economic evaluation in both the UK and Australia will comply with CHEERS 2022 recommendations(39).

## Data Collection and Management

### Data Collection

Participating sites will be expected to maintain a file of essential trial documentation, which will be provided by the CTRU, and keep copies of all completed paper CRFs. Participant questionnaires will be collected either on paper or electronically. Trial data collected on paper CRFs will be submitted to the CTRU usually via standard post or secure electronic transfer. All other data collection will be via Remote Data Entry (RDE) on electronic case report forms (eCRFs) onto a trial specific database managed by the CTRU at the University of Leeds. Data will only be completed by personnel authorised to do so by the PI and recorded on the trial-specific Authorised Personnel log.

### Confidentiality

Participant data collected during the trial will be kept strictly confidential and accessed only by delegated members of the research team. Copies of the UK participant consent forms will include participant names. Postal address, email address and phone number will also be collected depending on how a participant opts to complete their follow up. All other data collection forms that are transferred to or from the CTRU will be coded with a unique participant trial number. Data will be held securely on paper and electronically at the CTRU for the duration of the trial. The CTRU will have access to the entire database for monitoring, co-ordinating, and analysis purposes.

If a participant withdraws consent from further collection of data, their data collected up to the date of withdrawal will remain on file and will be included in the final trial analysis. Trial data will be retained for a minimum of 5 years. Data will be made available for secondary research once the main trial objectives are complete.

### Trial Management and Monitoring

Trial supervision will be established according to the principles of GCP and in line with the NHS UK Policy Framework for Health and Social Care. This includes a Trial Management Group (TMG), an independent Trial Steering Committee (TSC) and an independent Data Monitoring and Ethics Committee (DMEC).

The Trial Management Group, comprising the CI, CTRU team, CEDAR team, NHMRC CTC representatives, Australian Co-Chairs and other key external members of staff involved in the trial, and patient representatives have responsibility for the clinical set-up, on-going management, promotion of the trial, and for the interpretation of results. The DMEC is appointed to review the safety and ethics of the trial, alongside trial progress and the overall direction overseen by the TSC. Trial progress will be closely monitored by the DMEC by detailed unblinded reports prepared by the CTRU. The independent TSC will provide overall supervision of the trial.

The trial includes an internal pilot phase, within the first 12 months of open recruitment. The internal pilot will assess recruitment, sites and qualitative evaluations. Following the 12-month internal pilot the DMEC and TSC will review trial progress to independently advise on progression of the trial to the funder.

### Safety Reporting

An “Adverse Event” in POLARIS is defined as an untoward medical event in a participant which is related to the treatment of LARS. Examples of expected AEs in POLARiS include; wound-infection after SNM, pain or bleeding as a result of TAI and side effects of medications used for SNM. Adverse events will be graded for severity using CTCAE grading for all adverse events. Intra-operative adverse events will be graded using ClassIntra classification and post operative adverse events will be graded using the Clavien-Dindo Classification scale. Any serious AEs or related unexpected serious AEs should be reported to the CTRU within 24 hours. It is the role of either the PI or CI to assign relatedness and expected nature of serious AEs. Both the Data Monitoring & Ethics Committee (DMEC) and Trial Steering Committee perform periodic review of the safety data.

## Ethics and Dissemination

The trial will be performed in accordance with the principles of Good Clinical Practice and the Declaration of Helsinki (2013). This trial was reviewed and approved by Wales REC 4 (reference: 23/WA/0171) in the UK and Sydney Local Health District HREC (reference: 2023/ETH00749) in Australia. The appropriate local approval will be obtained by each participating site prior to entering participants into the trial. Further ethical approval of amendments to the protocol and/or related documents will be sought as necessary.

The trial outcomes will be disseminated to participants upon request and published on completion of the trial in a peer-reviewed journal and at international conferences. Credit for the main results will be given to all those who have collaborated in the trial. Authorship for the publication of the results of this study will be based on the principles of the International Committee of Medical Journal Editors Recommendation 2018. Data will be made available for secondary research once the main trial objectives are complete.

## Supporting information

Supplementary 1 Consent Form

Supplementary 2 Interview Guide

Supplementary 3 Interview Guide

## Data Availability

All data produced in the present study are available upon reasonable request to the authors.

