## Supplementary 1 Consent Form for "Pathway of Low Anterior Resection Syndrome (LARS) Relief After Surgery (POLARiS) Trial Protocol A prospective, international, open-label, multi-arm, phase 3 randomised superiority trial within a cohort, with economic evaluation, process evaluation and qualitative sub-study, to explore the natural hi"

Delete this line, then print on Trust/Hospital headed paper

| | |
|---|---|
| Participant ID: | Initials: |
| Date of Birth: | NHS/CHI Number: |
| ISRCTN: 12834598 | Principal Investigator: |

POLARiS
Pathway of Low Anterior Resection Syndrome relief after Surgery

### PARTICIPANT CONSENT FORM - POLARiS COHORT

***Please initial each box***

1. I confirm that I have read and understand the information sheet for the above study and have had the opportunity to ask questions. ☐
2. I understand that my participation in this study is voluntary and that I am free to withdraw at any time without my medical care or legal rights being affected. I understand that even if I withdraw from the above study, the data collected from me will be used in analysing the results of the study. ☐
3. I understand that my healthcare records may be looked at by authorised individuals from the study team, regulatory bodies or Sponsor in order to check that the study is being carried out correctly. ☐
4. I agree to complete the Quality of Life questionnaires and understand that my full name and address, or my email address and/or telephone number if I choose to complete the questionnaires online, will be passed to the CTRU for the purpose of issuing these questionnaires. ☐
5. I understand that if during this study my clinical care team determine that I have lost my ability to make my own decisions, no further study intervention will be given. I agree that data collected up until this point will remain on file and will be included in the analysis. ☐

6. I understand that the information collected about me may be used to support other research in the future, and may be shared anonymously with other researchers.
7. I understand that data may be passed to other organisations participating in this study (possibly in other countries where the data protection standards and laws are different to the UK) to monitor the safety of the treatment I am receiving.
8. I agree to a copy of this Consent Form being sent to the CTRU.
9. I agree that my GP, or any other doctor treating me, will be notified of my participation in this study.
10. I agree to take part in the study.

**The following point is OPTIONAL.** Even if you agree to take part in this study, you do not have to agree to this section

Please tick

| | Yes | No |
|---|---|---|
| Would you like to receive a lay summary of the trial results when the study is completed?<br>*The summary will be sent using your preferred method of contact as indicated fc please let your research team know if your contact details change.* | | |

**Patient:**

Signature…………………………………………………………………………………

Name (block capitals)……………………………………………………………………

Date…………………………………………………………………………………………

**Investigator:**

I have explained the study to the above named patient and they have indicated their willingness to participate.

Signature………………………………………………………………………………

Name (block capitals)…………………………………………………………………

Date…………………………………………………………………………………………

Delete this line, then print on Trust/Hospital headed paper

| Participant ID: | Initials: |
|---|---|
| Date of Birth: | NHS/CHI Number: |
| ISRCTN: 12834598 | Principal Investigator: |

POLARiS
Pathway of Low Anterior Resection Syndrome relief after Surgery

#### PARTICIPANT CONSENT FORM - POLARiS RCT

***Please initial each box***

11. I confirm that I have read and understand the information sheet for the above study and have had the opportunity to ask questions.

12. I understand that my participation in this study is voluntary and that I am free to withdraw at any time without my medical care or legal rights being affected. I understand that even if I withdraw from the above study, the data collected from me will be used in analysing the results of the study.

13. I understand that my healthcare records may be looked at by authorised individuals from the study team, regulatory bodies or Sponsor in order to check that the study is being carried out correctly.

14. I agree to complete the Quality of Life questionnaires and understand that my full name and address, or my email address and/or telephone number if I choose to complete the

questionnaires online, will be passed to the CTRU for the purpose of issuing these questionnaires.

15. I understand that if during this study my clinical care team determine that I have lost my ability to make my own decisions, no further study intervention will be given. I agree that data collected up until this point will remain on file and will be included in the analysis.

16. I understand that the information collected about me may be used to support other research in the future, and may be shared anonymously with other researchers.

17. I understand that data may be passed to other organisations participating in this study (possibly in other countries where the data protection standards and laws are different to the UK) to monitor the safety of the treatment I am receiving.

18. I agree to a copy of this Consent Form being sent to the CTRU.

19. I agree that my GP, or any other doctor treating me, will be notified of my participation in this study.

20. I agree to take part in the study.

**The following points are OPTIONAL.** Even if you agree to take part in this study, you do not have to agree to this section

| | Please tick ✓<br>Yes | No |
|---|---|---|
| I give my permission for the research team in CEDAR (Cardiff & Vale University Health Board) to contact me regarding participation in a patient interview (via telephone/video call). I understand that my contact and demographic information will be passed to CEDAR from the CTRU for this purpose. | ☐ | ☐ |
| Would you like to receive a lay summary of the trial results when the study is completed?<br>*The summary will be sent using your preferred method of contact as indicated for receiving questionnaires, please let your research team know if your contact details change.* | ☐ | ☐ |

**Patient:**

Signature……………………………………………………………………………………

Name (block capitals)……………………………………………………..……………………

Date………………………………………………………..……………………………………

**Investigator:**

I have explained the study to the above named patient and they have indicated their willingness to participate.

Signature………………………………………..…………………………………………

Name (block capitals)……………………………………………………..…………………

Date…………………………………………………..…………………………………………

(1 copy for patient; 1 for the CTRU; 1 held in patient notes, original stored in Investigator Site File)
