## Supplementary 2 Interview Guide for "Pathway of Low Anterior Resection Syndrome (LARS) Relief After Surgery (POLARiS) Trial Protocol A prospective, international, open-label, multi-arm, phase 3 randomised superiority trial within a cohort, with economic evaluation, process evaluation and qualitative sub-study, to explore the natural hi"

### POLARiS Patient Interview Topic Guide 1

Please note that these guides only represent the main themes to be discussed with the participants. Prompts may be used to guide the conversation. Non-leading and general prompts will also be used, such as “Can you please tell me a little bit more about that?”.

**MAIN PARTICIPANT INTERVIEW 1 (as soon after randomisation as possible)**

***INTRODUCTION***

*My name is XX and I work for an NHS department called Cedar. We are working with Leeds University on the POLARiS trial looking into bowel symptoms following colorectal surgery. You agreed to be contacted in relation to the POLARIS trial, is that correct?*

*Thank you very much for agreeing to be contacted and take part in this interview. I will be asking questions to better understand how patients experience LARS symptoms and the impact this has on your quality of life up until now. There are no right or wrong answers to any of our questions, we are interested in your own experiences.*

*The interview should take approximately one hour depending on how much information you would like to share. With your permission, I would like to audio/video record the interview because I don’t want to miss any of your comments. All responses will be kept confidential. This means that your interview responses will be stored anonymously and will only be shared with research team members. We will ensure that any information we include in our report does not identify you as the respondent. You may decline to answer any question or stop the interview at any time and for any reason. Are there any questions about what I have just explained?*

*Are you happy to start the interview? Are you happy for me to start recording?*

***RAPPORT***

*Establish rapport by asking a general question such as:* “tell me about yourself” or “how are you feeling today?”

***QUESTIONS***

**Experience of LARS symptoms**

- You were recruited into this trial because you are getting symptoms since having surgery on your bowel. These symptoms are sometimes called LARS (lower anterior resection syndrome) are you familiar with this term and happy for me to use it throughout the interview?
- Can you tell me a little about your LARS symptoms? What kind of symptoms have you experienced?
- How has having LARS symptoms impacted your daily life?
  - *(prompt: work, hobbies and interests, relationships with partners, family or friends, general wellbeing)*
- Which symptoms impact most on your daily life?

**Management of LARS**

- In what ways have you managed your LARS symptoms in the past before taking part in this trial?
  - (*prompt: medications, underwear/pads, staying close to toilet, not going out as much)*

**Preconceptions and expectations of LARS treatment**

- I understand that since agreeing to take part in this clinical trial that you will have been told that you are going to receive <<insert treatment allocation>> , is that right?
- What is your understanding of this treatment?
- How do you feel about the treatment you will be getting? Are there things you like or don't like about the treatment you will be getting?
- Have you started any treatment as part of the clinical trial? Can you tell me about that?
    - *(prompt: ease of use, barriers to engagement, information accessibility, support from staff)*
- What do you hope will change after using your assigned treatment?

**Support and information**

- Do you feel like you've had the right information and support you need to understand the treatment?
- What worked well and worked less well in terms of the support you have been given so far?

***WRAP-UP***

Thank you for taking the time to talk to me today. Your comments will be really helpful for our study and we appreciate your help. Do you have anything else you which to add before I stop the recording and stop the interview?
