## Supplementary 3 Interview Guide for "Pathway of Low Anterior Resection Syndrome (LARS) Relief After Surgery (POLARiS) Trial Protocol A prospective, international, open-label, multi-arm, phase 3 randomised superiority trial within a cohort, with economic evaluation, process evaluation and qualitative sub-study, to explore the natural hi"

### POLARiS Patient Interview Topic Guides 2&3

Please note that these guides only represent the main themes to be discussed with the participants. Prompts may be used to guide the conversation. Non-leading and general prompts will also be used, such as "Can you please tell me a little bit more about that?".

**MAIN PARTICIPANT INTERVIEWS 2 AND 3 (3m after treatment start & end of follow-up)**

***INTRODUCTION***

*My name is XX and I work for an NHS department called Cedar. We are working with Leeds University on the POLARiS trial looking into bowel symptoms following colorectal surgery. You agreed to be contacted in relation to the POLARIS trial, is that correct?*

*Thank you very much for agreeing to be contacted and take part in this interview. I will be asking questions to better understand how patients experience LARS symptoms and the impact this has on your quality of life prior. There are no right or wrong answers to any of our questions, we are interested in your own experiences. We want to talk to you now that you've been part of the trial for a while. We want to know more about how the treatment you have received for your LARS has worked for you.*

*The interview should take approximately one hour depending on how much information you would like to share. With your permission, I would like to audio/video record the interview because I don't want to miss any of your comments. All responses will be kept confidential. This means that your de-identified interview responses will only be shared with research team members and we will ensure that any information we include in our report does not identify you as the respondent. You may decline to answer any question or stop the interview at any time and for any reason. Are there any questions about what I have just explained?*

*Are you happy to start the interview? Are you happy for me to start recording?*

***RAPPORT***

*Establish report by asking a general question such as:* "how are you feeling today?" or "how have you been since we last spoke?"

***QUESTIONS***

**Assigned intervention**

- Last time we spoke you told me that as part of the clinical trial you would be receiving <<insert assigned intervention>> is that right?
- Has there been any change to this? For example, have you been allocated to a different treatment?
    - If so, how did this change come about and why?
    - <<if not move to next question>>

**Experience of the intervention**

- How have you been getting on with the treatment you are currently getting as part of the trial?
    - *(prompt: ease of use, barriers to engagement, information accessibility, support from staff)*

- How do you use or experience the treatment <<specific wording depending on assigned intervention>>
- What do you like about the treatment?
- What do you not like about the treatment?
- Is the treatment something you would carry on using?

**Intervention-specific questions (optional depending on answers to the above)**

- TAI
  - What type or brand of irrigation system are you using?
  - How did you find learning how to use the irrigation system?
    - *Prompt: teaching session with nurse, follow-up review, information*
  - How have you decided how often to use the device?
    - *Prompt: thinking about frequency, strategies, do they just use it when going out or for particular occasions or is it a habitual, every day activity*
- SNM
  - How did you find the procedures for having the electrodes implanted and then the stimulator 2 weeks later?
  - How have you found the device since it was implanted?
    - *Prompt: explanations beforehand sufficient, has it helped with symptoms, any issues with the implant*
- OCM
  - What advice were you given as part of receiving the OCM treatment?
    - *Prompt: diet, meds, physiotherapy*
  - Have you changed anything about the way you live your life as a result of the advice?
  - Did you receive enough support in order to make these changes?
  - Did you receive any other advice or support during this time?
    - *Prompt: referral to physiotherapy*
  - Is there anything you can think of that future patients might benefit from on the OCM pathway?

**Impact of intervention on LARS and quality of life**

- Has there been a change in your LARS symptoms since starting the treatment?
  - *(prompt: positive and negative changes relating to quality of life, symptoms, physical and psychological functioning)*
- How has this change come about?
- How have these changes, if any, impacted your daily life?
  - *(prompt: work, hobbies and interests, relationships with partners, family or friends, general wellbeing)*
- Have there been any other changes to your LARS symptoms during this time which you do not think are due to study treatment?

**Issues with the intervention**

- Have you experienced any problems with the treatment?
- What (if anything) might your healthcare team or the study team have done that would have helped you have a more positive experience of the treatment in the trial?
  - *(prompt: anything outside of your healthcare or study team)*

**Support and information**

- Have you had the right support to help you with the treatment? E.g. doctor, nurse, written material, family etc.
- Is the support easy to access?

***WRAP-UP***

Thank you for taking the time to talk to me today. Your comments will be really helpful for our study and we appreciate your help. Do you have anything else you which to add before I stop the recording and stop the interview?
